# Modeling cycle phases using hormone trajectories in women with and without polyendocrine metabolic ovarian syndrome

**DOI:** 10.64898/2026.06.02.26354701

**Authors:** Theresa M. Stujenske, Thomas P. Bouchard, Alexis Troy, Sara Kelemen, Brianna Folino, Tara Wills, Lauren A. Sugden

## Abstract

The recent availability of at-home menstrual cycle tracking technology has created opportunities for personalized assessment of reproductive health, alongside improved characterization of hormone patterns in women with and without reproductive disorders such as polyendocrine metabolic ovarian syndrome (PMOS), which affects approximately 10% of reproductive-age women. In this study, we leverage self-tracked urinary hormone data to develop an autoregressive Hidden Markov model (arHMM) that maps cycle days to physiologically meaningful phases based on hormone trajectories. By modeling day-to-day hormonal dynamics rather than absolute hormone levels, and allowing variable phase durations, this approach accommodates substantial variability in menstrual cycles, thereby enabling meaningful comparisons within and between individuals.

Across more than 3800 cycles from over 1100 individuals, we find that arHMM-derived phases reproduce expected hormonal patterns within follicular, periovulatory, and luteal phases, and that phase-based timing for hormone testing outperforms conventional cycle day–based testing in capturing the luteinizing hormone surge and post-ovulatory progesterone rise, highlighting limitations of fixed-day clinical protocols. We identify phase-specific differences between healthy controls and individuals with self-reported PMOS, including lower luteinizing hormone in the periovulatory phase, and reduced luteal-phase progesterone levels in PMOS. Furthermore, features derived from arHMM phase assignments enable classification of PMOS status with ∼78% accuracy, demonstrating the potential of this approach for non-invasive PMOS screening.

**Author Summary:** Tracking menstrual cycle information has become increasingly common, thanks in part to the availability of at-home urinary hormone data tracking technologies. In this study, we leveraged data from more than 3,800 menstrual cycles contributed by over 1,100 individuals in order to develop a model that predicts physiologically meaningful cycle phases based on day-to-day hormone patterns, and compared these patterns between individuals with and without polyendocrine metabolic ovarian syndrome (PMOS), a common endocrine disorder affecting approximately 1 in 10 reproductive age women. Our model successfully captures expected hormonal dynamics across six menstrual cycle phases and can be used to identify distinct phase-specific hormone patterns among individuals with PMOS. This study demonstrates the power of physiologically informed probabilistic modeling for leveraging data from at-home hormone monitoring towards a better characterization of menstrual cycle physiology and the development of noninvasive approaches for identifying reproductive health conditions such as PMOS.

## Introduction

Menstrual-related disorders affect approximately 10% of reproductive-age women, causing significant disease burden (1). Persistent delays in the diagnosis of these disorders highlight the need for non-invasive biomarkers to improve early detection (2,3). Hormonal trajectories across the menstrual cycle provide insight into women’s overall health and may serve as potential biomarkers of menstrual-related disorders like polyendocrine metabolic ovarian syndrome (PMOS), previously named polycystic ovary syndrome (4). Recent advances in tracking technologies that quantitatively measure urinary reproductive hormones across the menstrual cycle have expanded our ability to non-invasively track menstrual hormonal trajectories. Prior to these technologies, it was neither feasible nor cost-effective to track quantitative hormonal patterns across the menstrual cycle (5). Urine hormone testing is a convenient, low cost and accurate way to measure reproductive hormone levels (6), and changes in reproductive hormone levels and their metabolites excreted in the urine accurately reflect underlying menstrual cycle physiology (7). These self-tracked menstrual cycle data can be leveraged to identify novel hormonal biomarkers to improve early detection of common menstrual-related disorders (8,9).

Among healthy women, there is considerable variation in hormone levels across the menstrual cycle (10,11). To leverage these data to identify novel biomarkers, there is a need for computational models that account for this variability while also deciphering unique features with the potential to predict underlying hormonal disorders. Models based on average menstrual cycle parameters are inaccurate for predicting menstrual cycle phases in women with irregular cycles, a characteristic clinical feature of PMOS (12). Hidden Markov Models (HMMs), statistical frameworks widely used to model biological sequence data, are appropriate for labeling self-tracked time series due to their ability to describe complex relationships between observable and hidden biological states (i.e., phases of the menstrual cycle) (13–15). HMMs are well-suited to menstrual cycle data because they capture the underlying temporal dynamics that characterize hormonal changes across the cycle (16). Several studies have utilized HMMs to estimate ovulation timing and menstrual cycle phase with self-tracked data, including cervical fluid observations and basal body temperature (14,15) . To date, the HMM framework has not been used to predict menstrual cycle phase using self-tracked urinary reproductive hormone measurements, which may offer a more precise approximation of menstrual cycle physiology (6). By defining phases probabilistically, the HMM approach aligns more closely with physiological processes that vary naturally across women and over time. This flexibility allows for accurate modeling of hormonal fluctuations regardless of menstrual cycle length or ovulatory status. ***In this study, we use auto-regressive Hidden Markov Modeling (arHMM) to label menstrual hormonal trajectories utilizing self-tracked urinary reproductive hormone measurements. Using arHMM-derived features, we explore hormonal differences in women with and without self-reported PMOS and build a preliminary model for classifying women based on PMOS status*.**

## Materials and methods

This was a secondary data analysis of urinary hormone data prospectively collected using the Mira^TM^ Monitor (Quanovate Tech, Pleasanton, CA) in women with and without self-reported PMOS. Data included quantitative urinary reproductive hormone levels (estrone-3-glucuronide, E_1_3G, luteinizing hormone, LH and pregnanediol glucuronide, PDG), self-reported demographic information (date of birth), and self-reported health conditions (hormone imbalance, irregular cycle, PMOS, miscarriage history, recent birth control use, breastfeeding, postpartum not breastfeeding).

### Data Collection

Urinary hormone data was collected using the Mira monitor, an at-home hormone analyzer that syncs with the Mira App (Figure 1). The Mira monitor is an FDA and CE-registered fertility tracker used to measure urinary estrogen (E_1_3G), luteinizing hormone (LH) and urinary progesterone (PDG). When compared to the ClearBlue Fertility Monitor, an ultrasound-validated fertility monitor that identifies fertile days based on urinary E_1_3G and LH levels, the Mira monitor’s quantitative E_1_3G and LH levels correlated well with the ClearBlue monitor’s test results (8). The data is stored according to the consent given by users in the data use agreement. This study (protocol 2023/10/11) has been verified by the Institutional Review Board at Duquesne University as Exempt according to 45CFR46.101(b)(4): (4) Secondary Research Uses of Data or Specimens on 10/21/2023.

**FIGURE 1:**
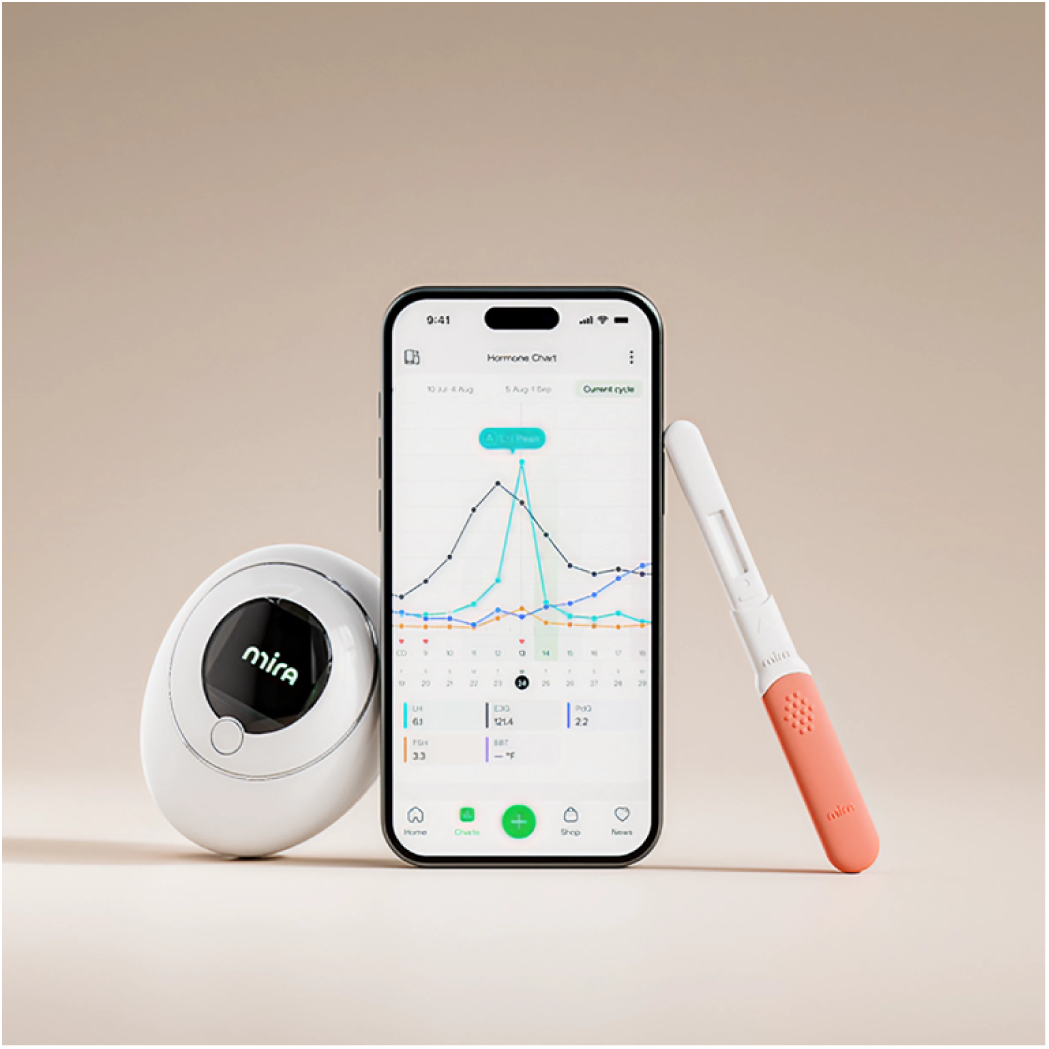
Mira^TM^ Monitor. *Image used with permission from Quanovate Tech Inc*.

Mira test wands detect urinary E_1_3G, LH and PDG levels based on fluorescence immunochromatography. The Mira monitor recognizes the test fluorescence signal and quantitatively converts it to the concentration of E_1_3G, LH and PDG. The user performs a test by dipping a Mira test wand into a urine sample for about 15 seconds and inserting it into the monitor. The results appear on the monitor in 21 minutes and can be synced to the Mira app through Bluetooth when paired with the monitor. The dataset included time-stamped urine hormone test results of E_1_3G, LH and PDG across recorded cycles.

The total dataset included 2,000 Mira users with at least three cycles of data, 1,000 Mira users with self-reported PMOS and 1,000 Mira users without self-reported PMOS. For all users, we removed cycles longer than 100 days as well as cycles recorded for users over the age of 45. We also implemented a filter to remove cycles without sufficient data using model-based criteria described below. The inclusion criteria for healthy controls were women between the ages of 18-45. Exclusion criteria were any self-reported conditions other than recently coming off birth control and miscarriage history. The inclusion criteria for participants with PMOS were women between the ages of 18-45 and self-reported PMOS. Exclusion criteria for all participants was self-reported breastfeeding. For subgroup analyses based on cycle lengths (cycle frequency), participants were defined as having normal cycle frequency if at least 90% of their cycles were between 24-38 days (17). Women with PMOS often have excessively long (>38 days) and irregular menstrual cycles (18). Since cycle length was the only phenotypic feature available in this dataset, we performed subgroup analyses to determine whether there were hormonal differences in healthy controls and women with self-reported PMOS based on cycle length.

The dataset included the following cycle-level information: cycle length, cycle start date, and cycle end date as well as all time-stamped urine hormone readings collected on a given cycle day. If a cycle day included more than one test result, we took the first recorded level of E_1_3G and PDG and the maximum LH level recorded that day. We aligned the cycle and hormone data in order to assign each hormone measurement to the corresponding day of the cycle (with day 1 representing the first day of bleeding), allowing for the visualization of single cycle patterns within and across individuals (“forward indexing”).

In order to account for variation in cycle length, and the increased variation of the follicular phase relative to the luteal phase (10), we also implemented two additional indexing strategies: 1) alignment to the end of the cycle (“backward indexing”), in which day -1 corresponds to the last day of the cycle, and 2) alignment to the LH surge (“LH-indexing”), in which day 0 corresponds to the day with the highest recorded LH value across that cycle, day -1 corresponds to the day prior to the LH surge, and day 1 corresponds to the day following the LH surge. These three indexing strategies allowed for the analysis and visualization of average hormonal trends within our healthy controls, with the average hormone patterns in the follicular phase, luteal phase, and around the LH surge brought into focus using forward-, backward- and LH-indexing respectively.

### Learning auto-regressive hormone patterns and building the autoregressive Hidden Markov model (arHMM)

To develop a “typical” model of day-to-day hormone patterns over the course of a single menstrual cycle, we first curated a set of cycles belonging to our healthy control population, filtering for cycles that were between 24 and 38 days long (17), for which missingness for each hormone (defined as the number of recordings divided by the cycle length) was less than 50% (testing on average every other day). We then identified six general phases based on the average hormone patterns represented via the three cycle indexing strategies described above, which together provided clearer patterns at the beginning and end of the cycle, as well as around the LH surge. Specifically, we divided the cycle into phases during which the directional trajectory of each hormone was consistent (i.e the hormone is rising, falling, or remaining constant throughout the phase). In our curated cycles, these phases approximately spanned the first 5 days of the cycle (phase 1), 10-2 days prior to the LH surge (phase 2), 2-0 days prior to the LH surge (phase 3), 0-4 days post LH surge (phase 4), 9-5 days before the end of the cycle (phase 5), and the last 5 days for the cycle (phase 6).

For each phase, and for each of the three hormones, we also built a simple linear regression model to predict the hormone level on day i+1 given the hormone level observed on day i, using all recorded consecutive days for which the second day belonged to the phase of interest (as specified in the previous paragraph). In order to enforce linear trajectories over multiple days, we set the slope to 1, and estimated the intercept (b^k_j) and variance (sigsq^k_j) for each hormone k and phase j=1,…,6. In practice, this captures the general behavior of each hormone in each phase: e.g. PDG tends to rise in phase 4 (positive intercept), while LH tends to fall (negative intercept), while allowing the absolute hormone levels on a given day to be variable within and between individuals.

The six cycle phases described above constitute the latent states in our autoregressive Hidden Markov model (arHMM) with a transition matrix that ensures that each cycle must progress through each phase in order, while allowing the number of days spent in each phase to vary. We set transition probabilities based on the observed number of days spent in each phase on average in our curated set of cycles (Supplementary Table 1).

The autoregressive piece of the arHMM comes from the emission model, which models the emission for a hormone k on day I, *X*_*ik* as a stochastic linear function of the latent state (phase) on day i (*Y*_*i*) and the hormone level on day i-1:

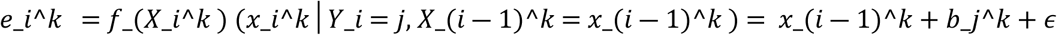

where *ϵ* ∼ *Normal*(0, *σ*_*j*^2*k*), and intercept *b*_*j*^*k* and variance *σ*_*j*^2*k* are the phase- and hormone-dependent constants learned from our data as described above (Supplementary Table 2). We make a simplifying assumption that the three hormones are conditionally independent given the latent states, such that the complete emission on day i is given by 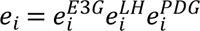.

**Table 2.**
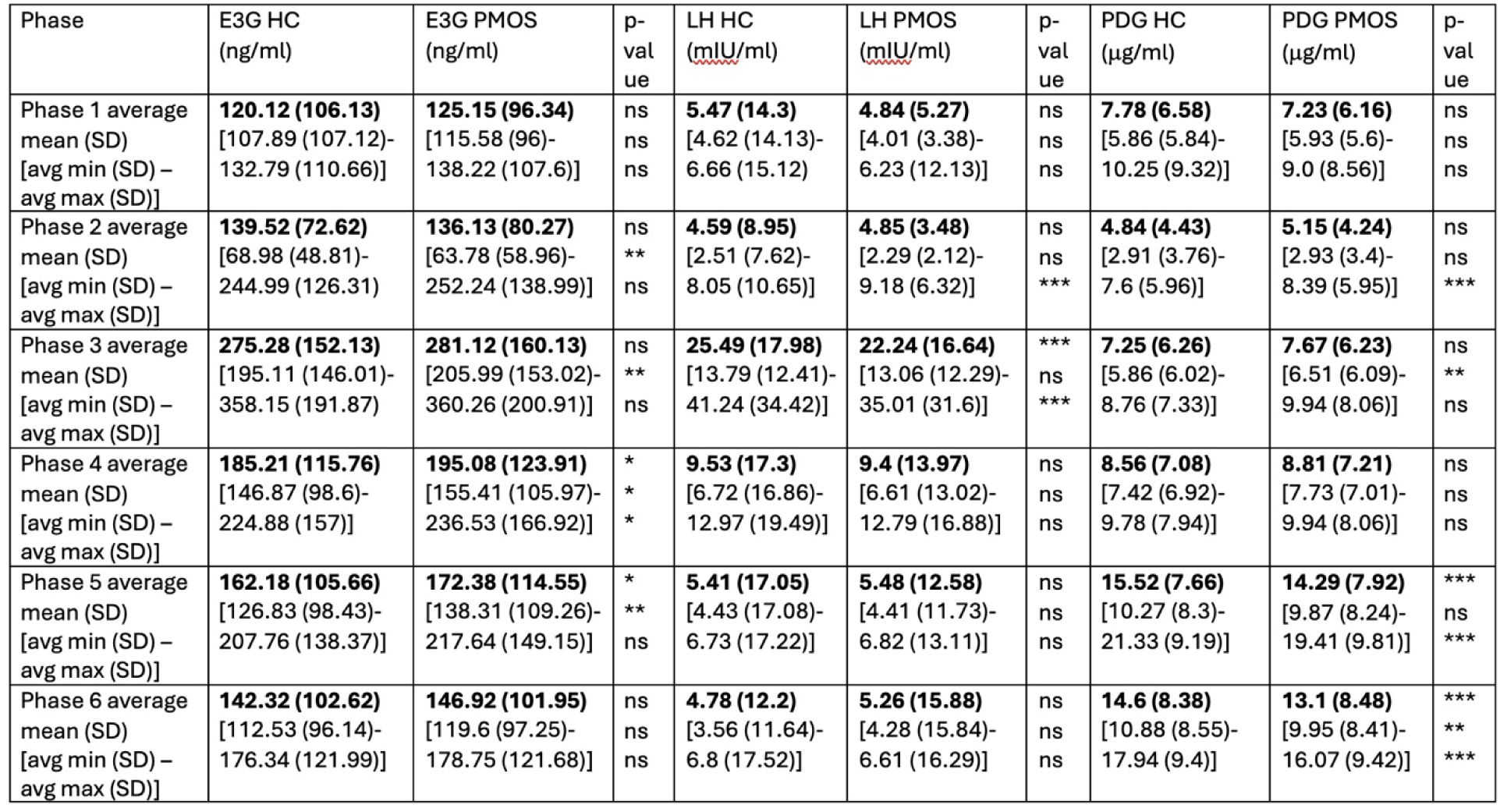
For each cycle, for each hormone and phase, we calculate the mean, minimum, and maximum for all available hormone measurements within that phase of the cycle (cycle-level features described in Methods). The table presents the averages and standard deviations of these cycle-level features separated into healthy control (HC) cycles and PMOS cycles. P-values are listed as not significant (ns), p<0.05 (*), p<0.01 (*), and p<0.001 (**).

We implemented a viterbi algorithm that finds the maximum *a posteriori* path through the latent states for a given cycle, using a start/end state to enforce that cycles begin by entering phase 1, and must exit from phase 6, having visited all phases in order in between. We also implemented a forward algorithm to calculate the log probability of the observed data, and as a basis for drawing an ensemble of representative phase paths for each cycle via stochastic backtrace. We used the log probability of the observed data as a measure of the fit of a cycle to the model.

To produce phase predictions for a cycle, that cycle must have exactly one recording each of PDG, E_1_3G, and LH for each day of the cycle. This required both imputing values for days on which the individual did not record their hormone data, and also reducing the amount of data on days when the individual recorded multiple times (as described above). To impute missing data, we used a linear interpolation approach: for any span of days with missing data, we took the nearest flanking hormone readings and assumed a linear trend in between. For missing data before the first recorded day, the hormone value was set to the value of the first available recording. For missing data after the last recorded day, the hormone value was set to the last available recording. Because of the importance of particular hormones in different phases of the menstrual cycle, we restricted all analyses to individuals with E_1_3G readings on at least 50% of phase 2 and phase 3 days, LH readings on at least 50% of phase 3 and phase 4 days, and PDG readings on at least 50% of phase 4 and phase 5 days.

### Exploring clinical applications

To compare hormone testing protocols using fixed cycle days vs arHMM-derived phases, we focused in particular on LH in phase 3 and on days 13-16 and on PDG in phase 5 and on days 19-22. For LH, we calculated the maximum hormone level for each cycle within the day or phase window, and for PDG, we calculated the average hormone level for each cycle within the day or phase window. We used paired t-tests to evaluate whether the hormone levels differed when considering phases vs days within each comparison.

To examine differences between healthy controls and women with self-reported PMOS, we developed individual-level features based on arHMM model output:

#### Hormone features

Within each cycle, for each hormone, and each HMM-predicted phase (1–6), we extracted the minimum, maximum, mean, median, and standard deviation of the hormone values that fall within that phase. This resulted in 90 cycle-level features. To construct individual-level features, we took the minimum, maximum, mean, median, and standard deviation of each cycle-level feature over all cycles the individual had recorded, yielding 450 individual-level features.

#### Phase length features

Within each cycle, we extracted the number of days spent in each of the six HMM-predicted phases, resulting in 6 cycle-level features. We then calculated the minimum, maximum, mean, median, and standard deviation for each phase length across all cycles the individual has recorded, yielding 30 individual-level features.

#### Model fit features

One output of the HMM is the joint log-probability of the entire sequence of hormones for a given cycle together with the maximum likelihood path, which can be interpreted as a measure of how well the cycle fit the model. From this single cycle-level feature, we generated five individual-level features by taking the minimum, maximum, mean, median, and standard deviation of the log-probability scores across all cycles recorded by an individual.

We constructed a random forest classifier using python library scikit-learn based on the arHMM-derived features described above (19), training the classifier to distinguish between individuals with PMOS from healthy controls. We also trained a second classifier that includes non-arHMM-derived cycle length features (mean, minimum, maximum, median, standard deviation, and range across all cycle lengths for the individual). Accuracy was calculated as the fraction of correct predictions pooled across true PMOS and healthy control individuals.

To label a cycle as ovulatory versus anovulatory, we built a proxy measure based on PDG levels in phase 5 (representing the post-ovulatory rise in progesterone). Average PDG levels in phase 3 for the same cycle are used as baseline PDG levels. For a cycle to be labeled ovulatory, we required a number of days (*n*) in phase 5 in which PDG levels were higher than the phase 3 baseline by a specific threshold. We considered thresholds ranging from a minimum of a 25% increase over phase 3 up to a maximum threshold of a 200% increase over phase 3, in increments of 25%. We considered a requirement of *n* = 1, 2, or 3 days. For cycles that did not have at least *n* PDG readings in phase 5, or had no recorded PDG readings in phase 3, we did not label the cycle as ovulatory or anovulatory. The remaining cycles were labeled anovulatory.

## Results

### Descriptive Analyses

This secondary analysis included 1,269 cycles from 277 healthy controls and 2,563 cycles from 841 women with self-reported PMOS (after applying inclusion and exclusion criteria described above in methods). Examples of two recorded cycles are shown in Figure 2A, one for a healthy control individual, and one for an individual with self-reported PMOS. Table 1 includes mean ages at the time of recording cycle data, mean cycle lengths, and mean number of cycles recorded for healthy controls and women with self-reported PMOS. On average, women with self-reported PMOS had longer and more irregular cycles as expected based on the clinical characteristics of this condition (Figure 2B).

**FIGURE 2:**
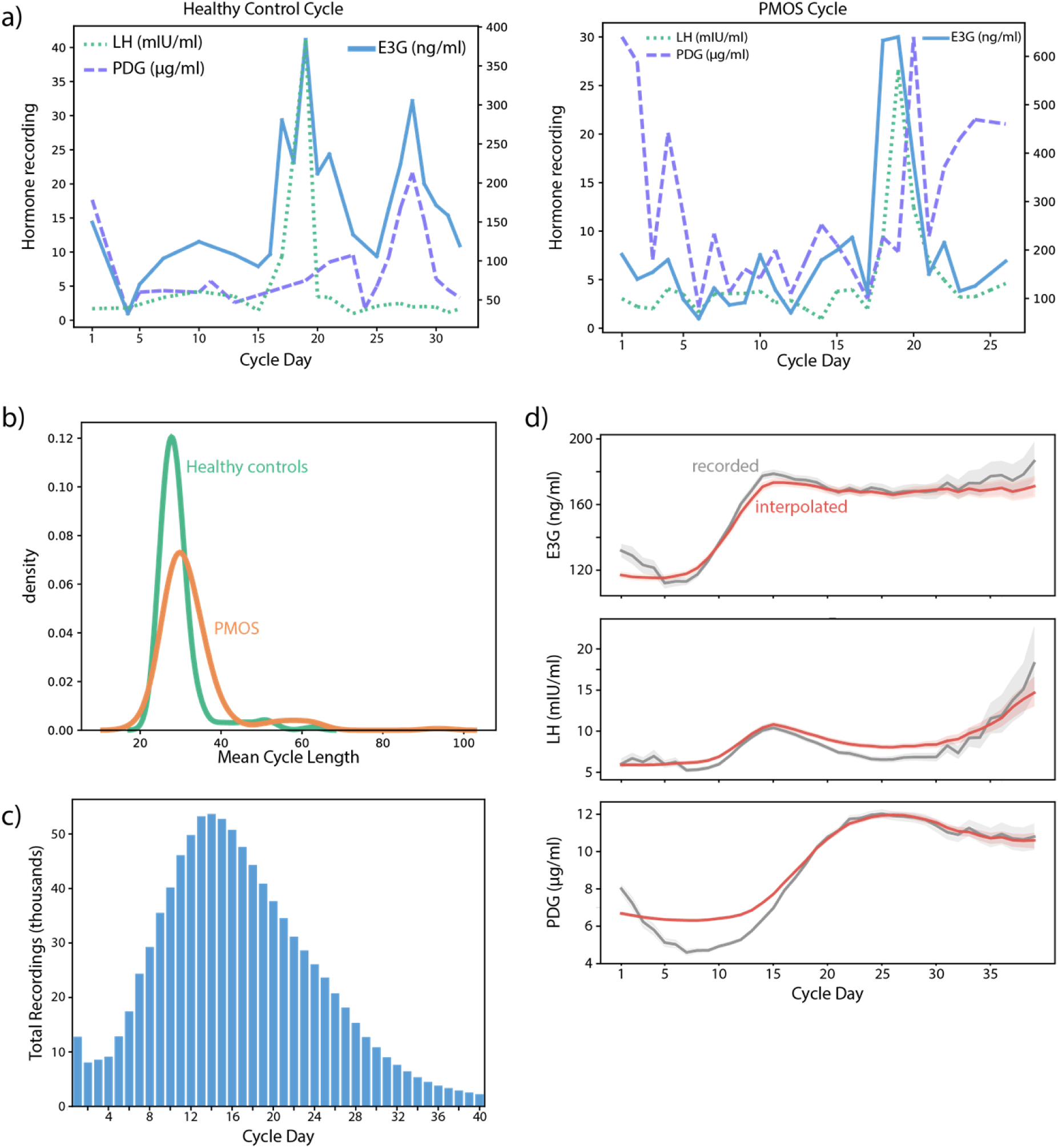
**A) Single cycle data**. LH, PDG, and E_1_3G recordings for two cycles, one belonging to a healthy control individual (left) and one belonging to an individual with self-reported PMOS (right). Hormone values for LH and PDG are on the left vertical axis, and values for E_1_3G are on the right vertical axis. **B) Distribution of cycle lengths.** Cycle lengths for healthy control cycles and PMOS cycles have overlapping distributions, but PMOS cycles were on average longer (mean of 28.55 days versus 32.05 days; p<0.001). **C) Recording behavior as a function of cycle day**. Individuals were much more likely to take hormone measurements towards the middle of the cycle, and there was a large amount of missing data at the beginning and end of the cycle. **D) Performance of missing data imputation**. In light of the missing data illustrated in panel C, we imputed recordings on missing days using the interpolation approach described in Methods. Gray lines represent available measurements for each hormone across cycle day, and red lines represent all measurements after interpolation. The interpolated values did not deviate dramatically from the available data, especially in realms where each hormone is dominant (E_1_3G in the luteal phase, LH around ovulation, and PDG in the follicular phase), though the approach did tend to overestimate PDG in the follicular phase.

**Table 1:** Study population characteristics. Age is recorded in years, and cycle length is recorded in days.

| Variable | Healthy Controls (n = 277) | PMOS (n = 841) |
| --- | --- | --- |
| Number of Cycles | 1269 | 2563 |
| Mean (SD) Age at Time of Recording Cycle | 36.3 (4.79) | 34.05 (4.44) |
| Mean (SD) Cycle Length | 28.55 (5.21) | 32.05 (7.55) |
| Mean (SD) Number of Cycles Recorded per Individual | 24.89 (11.28) | 20.11 (11.03) |
| % of Cycles Between 24-38 Days | 91% | 82% |

Hormone testing behavior was not uniform over the course of the menstrual cycle, with tests increasing towards the middle of recorded cycles around the presumed LH surge and ovulation (Figure 2C). Our analyses required imputation of missing data as described in Methods; we found that the imputed values tracked well with the available hormone levels in the dataset (Figure 2D). In particular, E_1_3G tracked well in the follicular phase when it is expected to be dominant, LH tracked well near the periovulatory window, and PDG tracked well in the luteal phase when it is expected to be dominant.

Absolute hormone levels across the menstrual cycle vary widely between individuals (10), which makes modeling these levels difficult with a standard HMM approach in which the model learns distributions for the absolute levels of each hormone throughout the cycle. To better suit this data and the way in which we expected hormone *trajectories* to shift over the different cycle phases, we implemented an autoregressive HMM (arHMM) which instead models the hormone levels as time series, with phases in which hormones are expected to rise, fall, or remain constant. As a result, two cycles with very different absolute hormone levels, but similar directional trajectories would be fit equally well by the model.

We used the three indexing strategies described in Methods to optimally map hormone levels to six cycle phases (1=early follicular, 2=late follicular, 3=periovulatory, 4=early luteal, 5=mid-luteal, and 6=late luteal) (Figure 3A-C).

**FIGURE 3:**
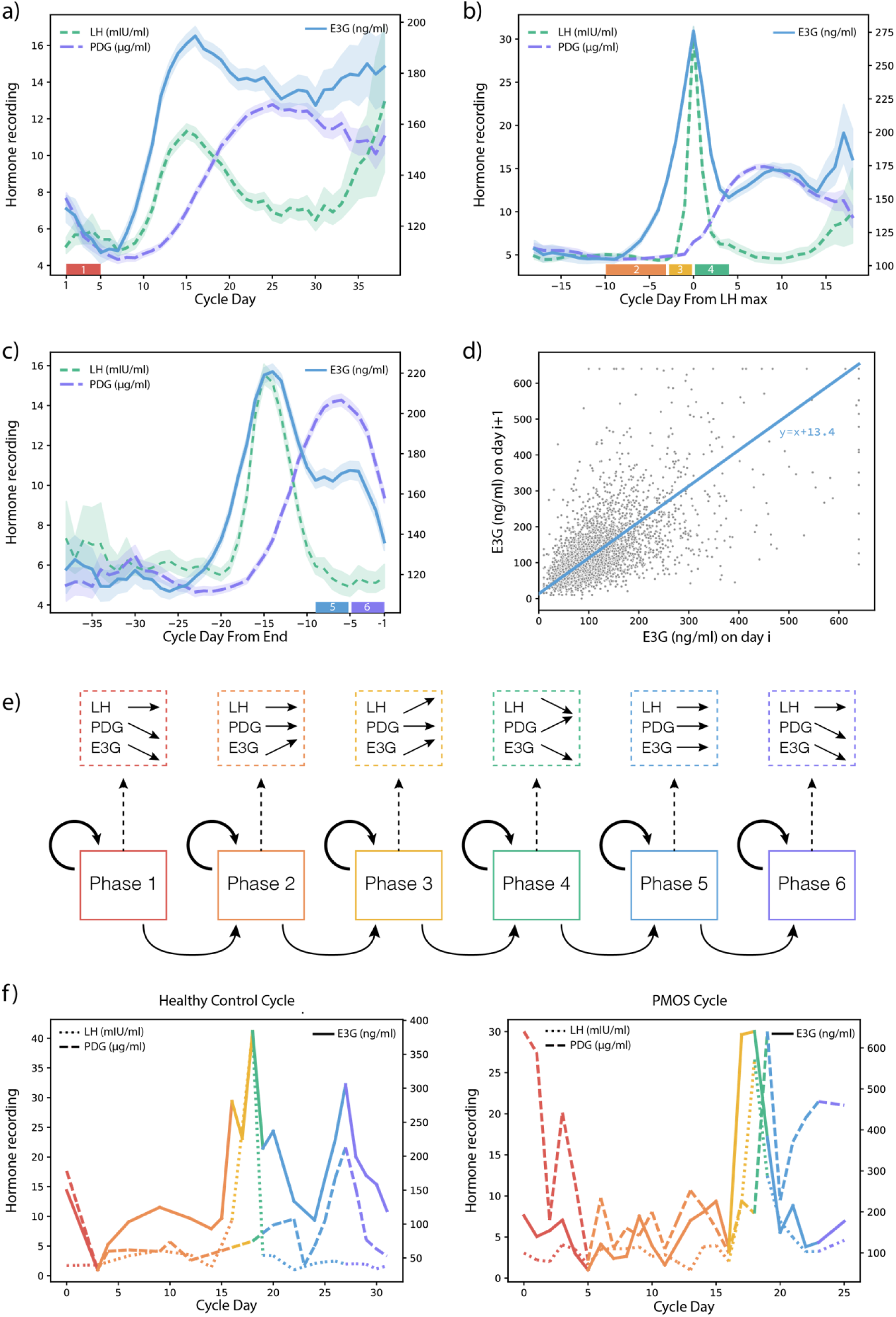
A-C) Forward, LH, and Backward indexing in cycles used for arHMM training. Forward indexing marks the first day of bleeding as day 1; this is used to learn trajectories in phase 1 (early follicular). LH indexing marks the day on which LH reaches a maximum as day 0; this is used to learn trajectories in phase 2 (late follicular), phase 3 (periovulatory), and phase 4 (early luteal). Backward indexing marks the last day of the cycle as day -1; this is used to learn trajectories in phase 5 (mid-luteal) and phase 6 (late luteal). **D) Learning phase trajectories.** E_1_3G hormone measurements are graphed for each pair of consecutive days in phase 2 within the curated set of cycles used for training the arHMM. In blue is the best fit line with a fixed slope of 1; within phase 2, we model E_1_3G rising by 13.4 ng/ml each day on average. **E) Schematic of arHMM.** Solid boxes outline the six hidden states of the arHMM, phases 1-6. Solid lines represent transition probabilities; paths through the model must start at phase 1 and end at phase 6 and can remain within phases for multiple days. Dotted boxes and lines represent the autoregressive emission model; within each phase, each hormone has an expected linear trajectory day over day (see Figure 3D). Briefly, PDG and E_1_3G are expected to fall in phase 1, E_1_3G rises in phase 2, E_1_3G and LH rise in phase 3, LH and E_1_3G fall in phase 4 while PDG rises, and PDG and E_1_3G fall in phase 6. **F) Example arHMM phase assignments.** For the same cycles as in Figure 2A, the arHMM viterbi path predicts phases for each day of the cycle, beginning in phase 1 (red), then moving in sequence through phase 2 (orange), phase 3 (yellow), phase 4 (green), phase 5 (blue), phase 6 (purple).

To develop the arHMM model, we used a curated set of 626 cycles from 189 healthy controls as described above, using these cycles to learn day-to-day trajectories for each hormone in each phase (Figure 3D and Supplementary Table 1). The full schematic of the arHMM model is included in Figure 3E. The arHMM model produces phase predictions for each cycle in the dataset (total cycles in each group included in Table 1); phase assignments for two example cycles are provided in Figure 3F.

We found that the average hormone levels within assigned phases across all cycles were consistent with expected menstrual cycle hormonal trajectories: E_1_3G rises in phase 2 and phase 3, LH surges in phase 3, and PDG rises in phases 4 and 5 (Figure 4A and Supplementary Figure 1).

**FIGURE 4:**
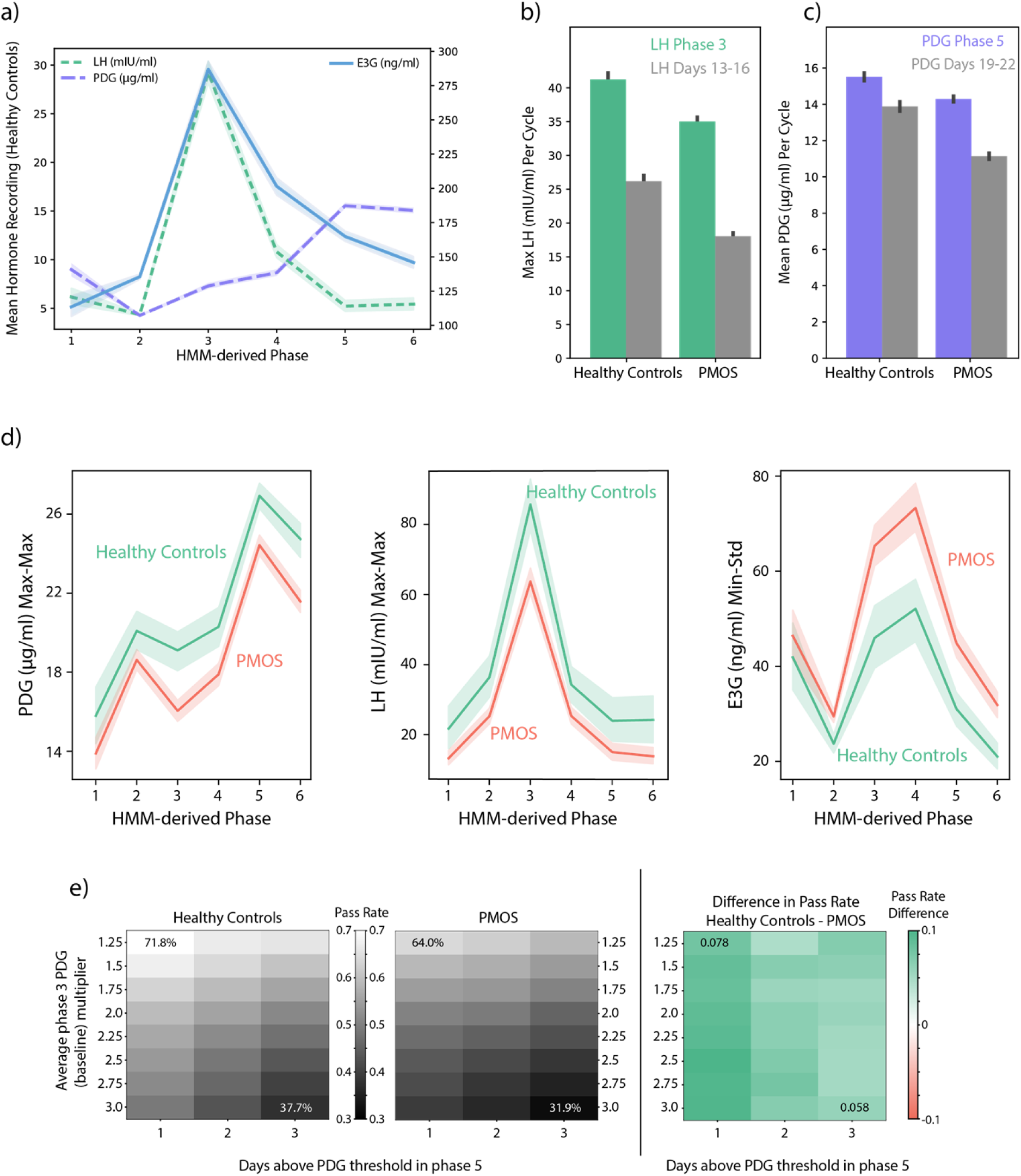
**A) Average hormone levels in each arHMM-derived phase**. Averaging over all healthy control cycles, and all recordings per phase within each cycle, the arHMM reproduced expected patterns: a rise in E_1_3G in phase 2 (late follicular) and phase 3 (ovulation), a rise in LH in phase 3 and subsequent fall in phase 4 (early luteal), and a rise in PDG in phase 5 (mid-luteal). **B) LH surges were significantly more robust in phase 3 than on days 13-16 of the menstrual cycle.** For each cycle, we calculated the maximum LH value observed in phase 3 and the maximum LH value observed across days 13-16. While PMOS cycles had lower LH values than healthy control cycles, within both groups, a cycle-day testing protocol is less effective at recording the LH surge than testing during phase 3. **C) PDG rises were significantly more robust in phase 5 than on days 19-22 of the menstrual cycle.** For each cycle, we calculated the mean PDG value observed in phase 5 and the mean observed across days 19-22. We saw a similar result; PMOS cycles have lower PDG levels than healthy control cycles on average, but within both groups, the cycle-day testing protocol is less effective at recording the luteal phase PDG rise. **D) arHMM-derived hormone features reveal differences in PMOS and healthy control individuals.** We calculated individual-level features as described in Methods; these aggregated hormone recordings across all available cycle days within a particular phase, followed by an aggregation across all cycles for an individual. Three examples of these individual level features are shown here, across all six phases. Left: Maximum PDG within each phase, maximized across all cycles; Middle: Maximum LH within each phase, maximized across all cycles; Right: Minimum E_1_3G within each phase, with standard deviation across all cycles. The latter reflects an increase in variability cycle-to-cycle within PMOS individuals. **E) Self-reported PMOS cycles were less likely to pass threshold-based ovulation tests.** Left (healthy controls) and middle (PMOS): heatmaps show the fraction of cycles that were labeled ovulatory for pairs of threshold parameters: a number of days (columns) with PDG levels over a particular value (rows; as a multiplier of average phase 3 PDG levels). Right: difference in pass rates, healthy controls - PMOS. Across the choices of threshold parameters, PMOS cycles were less likely to be labeled as ovulatory.

### Clinical applications of arHMM-derived phases

Clinicians order laboratory testing on specific days of the menstrual cycle to evaluate patients for underlying menstrual-hormonal abnormalities or to schedule ultrasounds for ovulation confirmation (20). For example, to evaluate periovulatory hormone patterns, laboratory testing may be ordered on cycle days 13-16. To examine the utility of testing hormone levels using arHMM-derived phases rather than fixed cycle days, we compared maximum LH levels in phase 3 to maximum LH levels on days 13-16. Hormonal testing is also frequently ordered on cycle days 19-22 to examine the post-ovulatory rise in progesterone as a useful proxy measure for ovulation (21). To examine the utility of phase-based testing, we compared PDG levels in phase 5 to mean PDG on days 19-22, as described in Methods. In both healthy controls and individuals with PMOS, we found that the phase-based levels are significantly higher than the cycle-day-based levels (p<0.001 for all four comparisons; Figure 4B-C). These findings suggest that laboratory testing on fixed cycle days may miss important physiological events, such as ovulation, due to the variability in timing of hormonal trajectories across the menstrual cycle (21).

Table 2 contains average cycle-level features (described in Methods) calculated within healthy control cycles and within PMOS cycles. We find that in many instances, hormone features within PMOS cycles differ significantly from those within healthy control cycles, including higher E_1_3G in phases 3, 4, and 5, lower LH in phase 3, and lower PDG in phases 5 and 6. In addition to analyzing mean hormone levels within phases, we also considered minimum and maximum levels within each phase, as a mechanism for examining potential differences in the hormone ranges (e.g. maximum LH in phase 3 tended to be higher in healthy controls compared to individuals with PMOS). We also found that the length of time spent in some phases differed significantly (Table 3); in particular, PMOS cycles spent a significantly longer time in phase 2 than healthy control cycles (p<0.001), which is consistent with the pathophysiology of PMOS, characterized by oligomenorrhea and ovulatory dysfunction (22). We also examined differences between subgroups based on cycle length (reported in Table 3 and Supplementary Tables 3 and 4). Overall, when grouped by cycle length (e.g. normal cycle length healthy controls vs normal cycle length PMOS) we still identified distinct hormonal patterns within the arHMM-derived cycle phases.

**Table 3:** The table presents the averages and standard deviations of the number of days spent in each phase separated into healthy control (HC) cycles and PMOS cycles and subgroups based on cycle length (normal cycle length vs. abnormal cycle length). P-values are listed as not significant (ns), p<0.05 (*), p<0.01 (*), and p<0.001 (**).

| arHMM-Phase | HC | PMOS | p-<br>value | HC<br>Normal Cycle<br>Length | PMOS<br>Normal<br>Cycle Length | p-<br>value | HC<br>Abnormal<br>Cycle Length | PMOS<br>Abnormal<br>Cycle Length | p-<br>value |
| --- | --- | --- | --- | --- | --- | --- | --- | --- | --- |
| Phase 1 | 1.34 (1.75) | 1.32 (1.98) | ns | 1.23 (1.37) | 1.27 (1.51) | ns | 1.47 (2.11) | 1.36 (2.23) | ns |
| Phase 2 | 13 (4.74) | 16.87 (7.81) | *** | 13.07 (4.18) | 14.4 (4.68) | *** | 12.92 (5.36) | 18.44 (8.92) | *** |
| Phase 3 | 1.82 (1.91) | 1.74 (1.66) | ns | 1.81 (1.76) | 1.75 (1.53) | ns | 1.82 (2.08) | 1.73 (1.73) | ns |
| Phase 4 | 1.51 (0.76) | 1.58 (1.11) | * | 1.5 (0.71) | 1.6 (1.05) | * | 1.51 (0.83) | 1.57 (1.14) | ns |
| Phase 5 | 3.63 (2.71) | 3.56 (3.06) | ns | 3.66 (2.69) | 3.53 (2.78) | ns | 3.6 (2.74) | 3.58 (3.23) | ns |
| Phase 6 | 7.25 (4.64) | 6.98 (4.5) | ns | 7.38 (4.53) | 7.11 (3.78) | ns | 7.1 (4.78) | 6.89 (4.91) | ns |

As described in Methods, we also aggregated cycle-level features into individual-level features that incorporated the arHMM output across all recorded cycles for a given individual. Figure 4D illustrates some of these individual-level features: we found that the hormone levels for E_1_3G, LH, and PDG had distinct patterns across the six phases for individuals with self-reported PMOS compared to healthy control individuals.

To explore the potential of these features to detect PMOS in individuals, we developed a preliminary random forest classifier as described in Methods. When using arHMM-derived hormone and phase length features, we achieved 78.4% accuracy at distinguishing between individuals with self-reported PMOS and healthy control individuals. If we included cycle length features, this accuracy increased slightly to 79.2%.

In examining the model fit features described in Methods, we observed that model fit declined as a function of cycle length, which is an expected pattern. When accounting for this relationship with cycle length, however, we do not see a difference in model fit between healthy control cycles and PMOS cycles (Supplementary Figure 2). This is likely a reflection of the flexibility of the arHMM itself, and its reliance on directional trajectories rather than absolute hormone levels; we were able to model PMOS cycles as effectively as healthy control cycles.

Previous studies suggest that a useful proxy measure of ovulation is when an LH surge is followed by elevated PDG levels for multiple days (21). As described in Methods, we labeled a cycle as ovulatory versus anovulatory based on PDG levels in phase 5 (representing the post-ovulatory rise in progesterone). Overall, cycles in women with PMOS were less likely to be labeled ovulatory based on these thresholds, in line with the expectation that women with PMOS have fewer ovulatory cycles (23) (Figure 4E).

## Discussion

In this study, we demonstrated that arHMMs offer a novel biologically grounded approach for modeling the menstrual cycle using quantitative self-tracked urinary reproductive hormone data. HMMs provide a computational framework for modeling menstrual-hormonal physiology, allowing for variability in cycle length and in the timing of key physiological events across the cycle. The autoregressive component allows the model to leverage day-to-day trajectories within each cycle phase (e.g. rising or falling) rather than relying on absolute hormone values, which are variable both within and between women.

We further demonstrated that arHMM-derived features revealed distinct differences in hormonal trajectories between healthy controls and women with self-reported PMOS. Specifically, women with PMOS exhibited lower LH levels in phase 3 (periovulatory phase), and lower PDG levels in Phases 5 and 6 (mid-luteal and late luteal phases). While previous studies have observed higher LH levels during the follicular phase in women with PMOS relative to healthy controls (which we also observed, though non-significant), the lower average LH levels in Phase 3 that we observed in women with PMOS may reflect an underlying dysregulation in hormonal feedback as a result of poor follicle development and anovulatory hormonal patterns (24–26). E_1_3G was also on average higher across the phases in women with self-reported PMOS, though these patterns were less clear, likely reflecting the heterogeneity of PMOS phenotypes present in this population (27,28). The recent name change more accurately reflects the multisystem pathophysiology of PMOS, with broad clinical features including reproductive, metabolic, psychological and dermatological features (4). To further develop a classifier for PMOS prediction based on arHMM-derived hormonal features, there is a need for future studies with well-defined populations of women with precise characterization of PMOS phenotypes.

Finally, we explored several clinical applications of arHMM-derived features. We demonstrated the potential utility of using arHMM-derived phase information compared to cycle day for scheduling hormonal testing to permit a more precise determination of hormonal levels based on underlying hormonal trajectories. Using arHMM-derived passed information may allow clinicians to provide a more personalized assessment of underlying hormonal health, especially for individuals with atypical or long cycles (29). We also examined the potential of arHMM-derived phase information to estimate a cycle’s ovulatory status based on PDG levels after the LH surge. With further research, this approach may offer a non-invasive means of determining whether an individual is experiencing anovulatory cycles. As ovulatory status is the most accurate reflection of underlying menstrual-hormonal health (30), there is a critical need for developing accessible and non-invasive methods for evaluating menstrual health in an ongoing and dynamic way (9). Further development of this approach also holds the potential of providing real-time feedback to individuals based on self-tracked hormonal data.

The arHMM model also has important applications for menstrual cycle research. There is no universally accepted method for measuring the menstrual cycle as a continuous variable (31). Cycle-day-based approaches tend to oversimplify underlying hormonal dynamics and fail to capture individual variability in ovulation timing, which can lead to misidentification of important physiological events. Recent studies have shown the utility of dividing the menstrual cycle into sub-phases as a means of providing greater insight into the underlying hormonal environment (31,32). The arHMM allows for identification of these latent sub-phases based on quantitative patterns and can be applied more broadly to map hormonal data to relevant menstrual-physiological states. Increasingly, menstrual phase is incorporated as an important predictor in biobehavioral research, creating a need for consistent and precise methods for determining menstrual phase, like the arHMM described here (5,33,34).

### Limitations

In this study, we demonstrated the potential of the arHMM framework for modeling menstrual cycle dynamics using self-tracked hormonal data. However, several limitations should be considered when interpreting the findings. First, there are currently no published ultrasound-validated reference ranges for the Mira monitor. Although prior research has shown strong correlation between the Mira monitor and the ultrasound-validated ClearBlue monitor (8), further validation against ultrasound-confirmed ovulation is needed before these findings can be more broadly generalized. Second, this study was a secondary analysis of self-tracked, real-time urinary hormone data. The observational nature of these data introduces challenges related to missingness and uneven sampling. Because data collection occurred outside of a controlled research setting, participants were more likely to record hormone levels during the mid-cycle period, when hormonal monitoring is most relevant for predicting and confirming ovulation. As a result, data from the early follicular and late luteal phases were relatively sparse, limiting our ability to draw robust conclusions about hormonal patterns during these time points.

Additionally, diagnostic information, including PMOS, was self-reported. While self-report may be considered a reasonable approach in population-based research, it limits the ability to examine distinct PMOS phenotypes (35). Finally, the study population may not be representative of the general population. Individuals who use fertility monitors are more likely to be actively trying to conceive, making it challenging to identify a healthy control group. To address these limitations, future prospective studies with well-defined populations, standardized data collection, and ultrasound-validated hormone ranges are needed.

## Conclusion

The integration of probabilistic modeling with home-based quantitative hormone tracking represents an important step toward scalable, data-driven menstrual health assessment. Home-based devices capable of quantifying urinary E_1_3G, LH and PDG metabolites enable frequent, high-resolution sampling that captures intra- and inter-cycle variability. When paired with the arHMM framework, these data provide individualized insight into hormonal dynamics, offering a window into reproductive physiology that is both accessible and physiologically meaningful. Such approaches may facilitate longitudinal tracking of hormonal health, helping women and clinicians identify changes in ovulatory function or signs of endocrine dysfunction over time. Although additional validation in larger and more diverse cohorts is needed, the findings highlight the potential of combining home-based hormonal testing with advanced time-series modeling to reduce diagnostic delays, improve access to PMOS screening, and support personalized reproductive health monitoring.

## Code Availability

All code for model training and application, analysis, and generation of figures is available at https://github.com/lasugden/arHMM-cycle-tracking/

## Data Availability Statement

The data used in this study were obtained from a third-party source, Mira Fertility, and consist of de-identified menstrual cycle and urinary hormone monitoring data collected through the Mira Fertility platform and associated mobile application. The dataset includes user-collected reproductive hormone measurements collected using the Mira monitor. The authors obtained permission to use these data through a formal data use agreement with <u>Mira Fertility</u>.

Because the data are proprietary and subject to privacy, ethical, and contractual restrictions, the dataset is not publicly available.

Researchers interested in obtaining access to the data should contact <u>Mira</u> <u>Fertility</u> directly to inquire about data access policies, collaboration opportunities, and applicable data use agreements. Access may be restricted to qualified investigators and may require institutional review board (IRB) approval, execution of confidentiality agreements, and approval by the data provider. Known restrictions to data accessibility include proprietary ownership of the dataset, participant privacy protections, and limitations outlined in the governing data use agreement between the investigators and Mira Fertility.

## Acknowledgments

We would like to acknowledge Ilya Kuznetsov at Mira Fertility for providing access to the dataset used in this secondary data analysis. The authors thank the company and study participants whose contributions made this research possible. Mira had no role in the analysis, interpretation of findings, or decision to publish the results.

## Supplementary Materials

**Supplementary Table 1.** **HMM transition matrix:** Transition probabilities from row state to column state. The arHMM enforces movement from phase 1 through phase 6 in order.

|  | Start/End | Phase 1 | Phase 2 | Phase 3 | Phase 4 | Phase 5 | Phase 6 |
| --- | --- | --- | --- | --- | --- | --- | --- |
| Start/End | 0.0 | **1.0** | 0.0 | 0.0 | 0.0 | 0.0 | 0.0 |
| Phase 1 | 0.0 | **0.833** | **0.167** | 0.0 | 0.0 | 0.0 | 0.0 |
| Phase 2 | 0.0 | 0.0 | **0.888** | **0.112** | 0.0 | 0.0 | 0.0 |
| Phase 3 | 0.0 | 0.0 | 0.0 | **0.667** | **0.333** | 0.0 | 0.0 |
| Phase 4 | 0.0 | 0.0 | 0.0 | 0.0 | **0.8** | **0.2** | 0.0 |
| Phase 5 | 0.0 | 0.0 | 0.0 | 0.0 | 0.0 | **0.8** | **0.2** |
| Phase 6 | **0.167** | 0.0 | 0.0 | 0.0 | 0.0 | 0.0 | **0.833** |

**Supplementary Table 2.** **HMM Emission Parameters:** For each phase and each hormone, we enforce a slope of 1 (in order to maintain a linear trajectory over multiple days) and learn the intercept and standard deviation of the linear regression model. As an example, we expect an average day-over-day rise in LH of about 17 mIU/ml while in phase 3.

| **Phase** | **Hormone** | **Slope** | **Intercept** | **Std Dev** |
| --- | --- | --- | --- | --- |
| 1 | LH | **1** | **-0.343** | **6.563** |
| 2 | LH | **1** | **0.193** | **5.085** |
| 3 | LH | **1** | **17.024** | **24.123** |
| 4 | LH | **1** | **-9.341** | **11.151** |
| 5 | LH | **1** | **-0.308** | **7.587** |
| 6 | LH | **1** | **0.458** | **7.867** |
| 1 | E3G | **1** | **-0.551** | **64.526** |
| 2 | E3G | **1** | **12.912** | **76.119** |
| 3 | E3G | **1** | **43.739** | **125.849** |
| 4 | E3G | **1** | **-35.132** | **98.471** |
| 5 | E3G | **1** | **0.033** | **88.106** |
| 6 | E3G | **1** | **-11.458** | **84.215** |
| 1 | PDG | **1** | **-0.506** | **4.646** |
| 2 | PDG | **1** | **-0.015** | **3.987** |
| 3 | PDG | **1** | **0.787** | **4.451** |
| 4 | PDG | **1** | **1.658** | **6.296** |
| 5 | PDG | **1** | **0.529** | **7.649** |
| 6 | PDG | **1** | **-1.388** | **7.454** |

**Supplementary Table 3.** For each cycle, for each hormone and phase, we calculate the mean, minimum, and maximum for all available hormone measurements within that phase of the cycle (cycle-level features described in Methods). The table presents the averages and standard deviations of these cycle-level features separated into healthy control (HC) normal length cycles and PMOS normal length cycles (24-38 days). P-values are listed as not significant (ns), p<0.05 (*), p<0.01 (*), and p<0.001 (**).

| Phase | E3G HC<br>(ng/ml) | E3G PMOS<br>(ng/ml) | p-<br>val<br>ue | LH HC<br>(mIU/ml) | LH PMOS<br>(mIU/ml) | p-<br>val<br>ue | PDG HC<br>(µg/ml) | PDG PMOS<br>(µg/ml) | p-<br>val<br>ue |
| --- | --- | --- | --- | --- | --- | --- | --- | --- | --- |
| Phase 1 average<br>mean (SD)<br>[avg min (SD) –<br>avg max (SD)] | **112.8 (98.99)**<br>[102.3 (98.81)-<br>123.69 (103.78)] | **130.79 (95.84)**<br>[119.43 (95.76)-<br>144.01 (105.75)] | ns<br>ns<br>ns | **4.93 (6.27)**<br>[4.25 (6.14)-<br>5.85 (7.23)] | **5.21 (4.69)**<br>[4.42 (3.71)-<br>6.24 (6.78)] | ns<br>ns<br>ns | **7.68 (6.24)**<br>[6.0 (5.62)-<br>9.54 (8.5)] | **9.08 (6.45)**<br>[7.13 (5.94)-<br>11.37 (9.07)] | *<br>ns<br>ns |
| Phase 2 average<br>mean (SD)<br>[avg min (SD) –<br>avg max (SD)] | **138.89 (65.73)**<br>[69.2 (42.35)-<br>243.73 (121.6)] | **140.21 (79.61)**<br>[66.28 (55.9)-<br>255.74 (142.82)] | ns<br>ns<br>ns | **4.52 (9.84)**<br>[2.62 (9.86)-<br>7.84 (10.36)] | **4.41 (2.39)**<br>[2.13 (1.55)-<br>8.37 (4.91)] | ns<br>ns<br>ns | **4.61 (4.37)**<br>[2.81 (3.73)-<br>7.28 (5.95)] | **5.39 (4.39)**<br>[3.17 (3.53)-<br>8.46 (6.11)] | ***<br>ns<br>*** |
| Phase 3 average<br>mean (SD)<br>[avg min (SD) –<br>avg max (SD)] | **280.42 (154.71)**<br>[197.61 (148.53)-<br>366.26 (195.27)] | **291.38 (156.89)**<br>[208.87 (151.34)-<br>378.6 (197.99)] | ns<br>ns<br>ns | **26.21 (18.36)**<br>[13.46 (11.59)-<br>42.82 (32.84)] | **22.84 (15.36)**<br>[13.41 (11.89)-<br>35.9 (29.96)] | ***<br>ns<br>*** | **7.03 (6.26)**<br>[5.7 (6.1)-<br>8.54 (7.29)] | **7.95 (6.3)**<br>[6.75 (6.17)-<br>9.22 (7.16)] | **<br>**<br>ns |
| Phase 4 average<br>mean (SD)<br>[avg min (SD) –<br>avg max (SD)] | **183.12 (110.91)**<br>[144.26 (90.75)-<br>223.75 (156.79)] | **197.31 (117.44)**<br>[154.2 (96.79)-<br>244.39 (169.08)] | *<br>*<br>* | **8.98 (11.06)**<br>[6.12 (10.24)-<br>12.47 (14.14)] | **9 (12.11)**<br>[6.32 (10.91)-<br>12.22 (14.81)] | ns<br>ns<br>ns | **8.29 (6.91)**<br>[7.19 (6.71)-<br>9.44 (7.77)] | **9.1 (7.16)**<br>[8.04 (6.98)-<br>10.23 (8.03)] | *<br>*<br>* |
| Phase 5 average<br>mean (SD)<br>[avg min (SD) –<br>avg max (SD)] | **154.06 (86.66)**<br>[119.08 (79.46)-<br>200.59 (125.12)] | **169.22 (102.86)**<br>[135.34 (98.3)-<br>211.01 (137.54)] | **<br>***<br>ns | **4.63 (8.08)**<br>[3.59 (8.08)-<br>6.01 (8.58)] | **4.81 (8.73)**<br>[3.83 (8.4)-<br>6.07 (9.36)] | ns<br>ns<br>ns | **15.38 (7.41)**<br>[10.05 (8.09)-<br>21.34 (9.06)] | **15.36 (7.57)**<br>[10.63 (8.39)-<br>20.65 (9.04)] | ns<br>ns<br>ns |
| Phase 6 average<br>mean (SD)<br>[avg min (SD) –<br>avg max (SD)] | **134.86 (87.36)**<br>[107.09 (83.22)-<br>166.77 (107.59)] | **138.79 (89.51)**<br>[110.41 (85.23)-<br>171.16 (109.37)] | ns<br>ns<br>ns | **4.48 (8.52)**<br>[3.25 (7.83)-<br>6.48 (15.13)] | **4.73 (14.21)**<br>[3.7 (14.1)-<br>6.22 (14.88)] | ns<br>ns<br>ns | **14.58 (8.18)**<br>[10.66 (8.32)-<br>17.94 (9.34)] | **14.45 (8.45)**<br>[11.22 (8.73)-<br>17.21 (9.06)] | ns<br>ns<br>ns |

**Supplementary Table 4.** For each cycle, for each hormone and phase, we calculate the mean, minimum, and maximum for all available hormone measurements within that phase of the cycle (cycle-level features described in Methods). The table presents the averages and standard deviations of these cycle-level features separated into healthy control (HC) abnormal length cycles and PMOS abnormal length cycles. P-values are listed as not significant (ns), p<0.05 (*), p<0.01 (*), and p<0.001 (**).

| Phase | E3G HC<br>(ng/ml) | E3G PMOS<br>(ng/ml) | p-<br>val<br>ue | LH HC<br>(mIU/ml) | LH PMOS<br>(mIU/ml) | p-<br>val<br>ue | PDG HC<br>(µg/ml) | PDG PMOS<br>(µg/ml) | p-<br>val<br>ue |
| --- | --- | --- | --- | --- | --- | --- | --- | --- | --- |
| Phase 1 average<br>mean (SD)<br>[avg min (SD) –<br>avg max (SD)] | **126.18 (111.61)**<br>[112.51 (113.61)-<br>140.31 (115.79)] | **122.59 (96.57)**<br>[113.83 (96.17)-<br>135.59 (108.44)] | ns<br>ns<br>ns | **5.91 (18.49)**<br>[4.92 (18.29)-<br>7.32 (19.36)] | **4.67 (5.51)**<br>[3.83 (3.21)-<br>6.22 (13.9)] | ns<br>ns<br>ns | **7.89 (6.9)**<br>[5.72 (6.04)-<br>10.92 (10.02)] | **6.32 (5.81)**<br>[5.34 (5.33)-<br>7.83 (8.05)] | *<br>ns<br>*** |
| Phase 2 average<br>mean (SD)<br>[avg min (SD) –<br>avg max (SD)] | **140.29 (80.34)**<br>[68.71 (55.79)-<br>246.55 (131.97)] | **133.53 (80.6)**<br>[62.19 (60.8)-<br>250.02 (136.5)] | ns<br>*<br>ns | **4.69 (7.71)**<br>[2.37 (3.18)-<br>8.31 (11)] | **5.14 (4)**<br>[2.39 (2.41)-<br>9.7 (7.03)] | ns<br>ns<br>** | **5.12 (4.49)**<br>[3.05 (3.79)-<br>8 (5.94)] | **5.01 (4.13)**<br>[2.78 (3.32)-<br>8.35 (5.85)] | ns<br>ns<br>ns |
| Phase 3 average<br>mean (SD)<br>[avg min (SD) –<br>avg max (SD)] | **268.94 (148.78)**<br>[192.04 (142.93)-<br>348.17 (187.29)] | **274.59 (161.87)**<br>[204.16 (154.11)-<br>348.62 (201.95)] | ns<br>ns<br>ns | **24.61 (17.49)**<br>[14.2 (13.34)-<br>39.3 (36.22)] | **21.87 (17.4)**<br>[12.84 (12.54)-<br>34.45 (32.59)] | **<br>*<br>** | **7.53 (6.25)**<br>[6.07 (5.92)-<br>9.02 (7.37)] | **7.48 (6.18)**<br>[6.36 (6.03)-<br>8.8 (7.13)] | ns<br>ns<br>ns |
| Phase 4 average<br>mean (SD)<br>[avg min (SD) –<br>avg max (SD)] | **187.74 (121.44)**<br>[150.04 (107.38)-<br>226.26 (157.38)] | **193.67 (127.86)**<br>[156.18 (111.45)-<br>231.52 (165.39)] | ns<br>ns<br>ns | **10.19 (22.65)**<br>[7.45 (22.4)-<br>13.57 (24.45)] | **9.65 (15.03)**<br>[6.79 (14.21)-<br>13.15 (18.06)] | ns<br>ns<br>ns | **8.89 (7.27)**<br>[7.7 (7.17)-<br>10.18 (8.12)] | **8.62 (7.23)**<br>[7.53 (7.02)-<br>9.75 (8.07)] | ns<br>ns<br>ns |
| Phase 5 average<br>mean (SD)<br>[avg min (SD) –<br>avg max (SD)] | **171.79 (123.87)**<br>[136.01 (116.4)-<br>216.25 (152.25)] | **175.01 (121.31)**<br>[140.18 (115.64)-<br>221.83 (155.94)] | ns<br>ns<br>ns | **6.35 (23.59)**<br>[5.43 (23.63)-<br>7.58 (23.66)] | **5.91 (14.48)**<br>[4.77 (13.4)-<br>7.29 (14.99)] | ns<br>ns<br>ns | **15.68 (7.95)**<br>[10.56 (8.55)-<br>21.31 (9.36)] | **13.61 (8.07)**<br>[9.38 (8.11)-<br>18.62<br>(10.19)] | ***<br>**<br>*** |
| Phase 6 average<br>mean (SD)<br>[avg min (SD) –<br>avg max (SD)] | **150.69 (116.96)**<br>[118.64 (108.62)-<br>187.09 (135.69)] | **151.42 (107.99)**<br>[124.68 (102.98)-<br>182.95 (127.83)] | ns<br>ns<br>ns | **5.12 (15.33)**<br>[3.9 (14.8)-<br>7.16 (19.89)] | **5.56 (16.73)**<br>[4.59 (16.73)-<br>6.83 (17.02)] | ns<br>ns<br>ns | **14.62 (8.62)**<br>[11.16 (8.81)-<br>17.94 (9.49)] | **12.25 (8.4)**<br>[9.15 (8.1)-<br>15.35 (9.58)] | ***<br>***<br>*** |

**Supplementary Figure 1.**
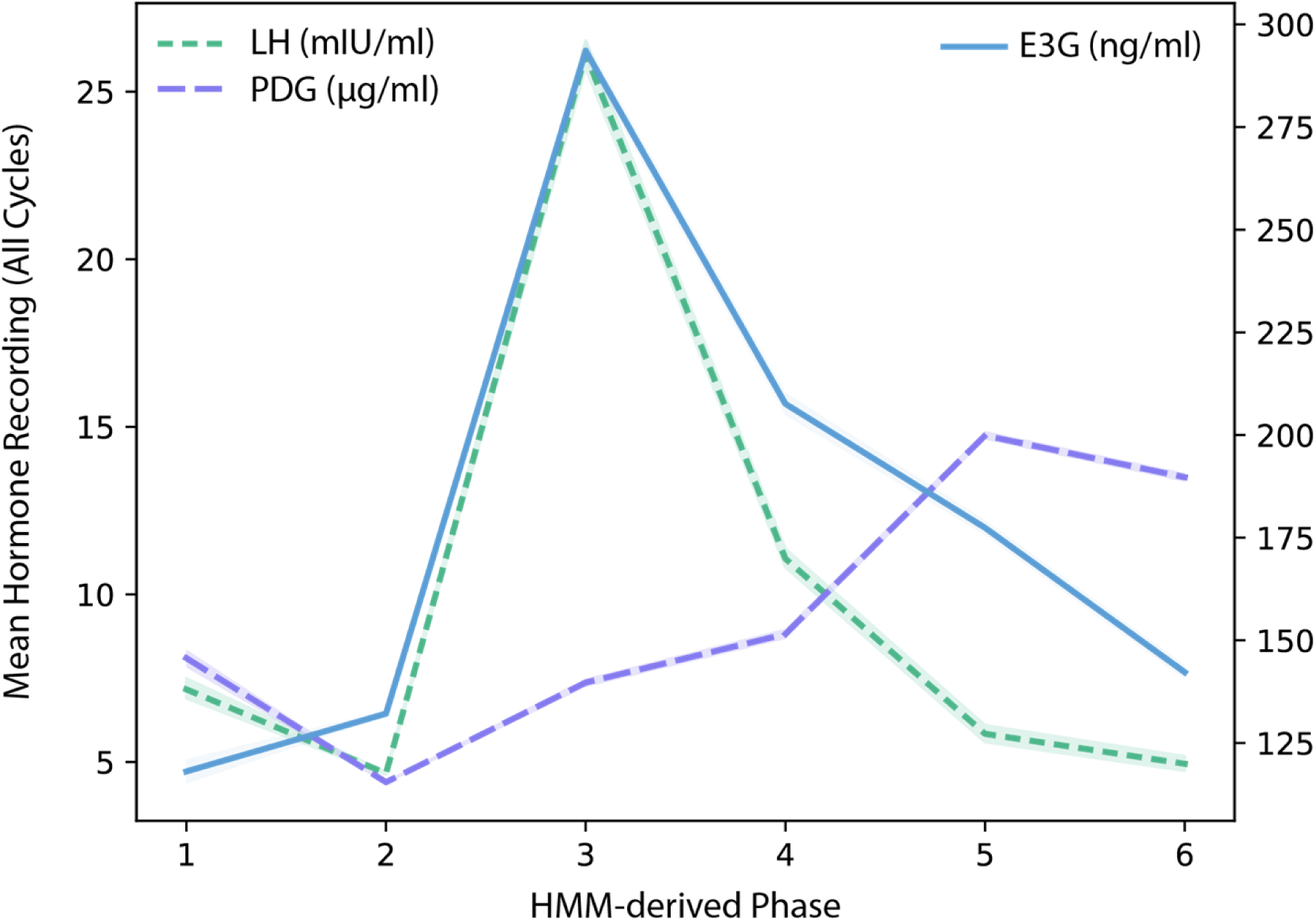
**Average hormone patterns over phases, all cycles:** Averaging over all healthy control cycles, and all recordings per phase within each cycle, the arHMM reproduces expected patterns: a rise in E3G in phase 2 (late follicular) and phase 3 (ovulation), a rise in LH in phase 3 and subsequent fall in phase 4 (early luteal), and a rise in PDG in phase 5 (mid-luteal).

**Supplementary Figure 2.**
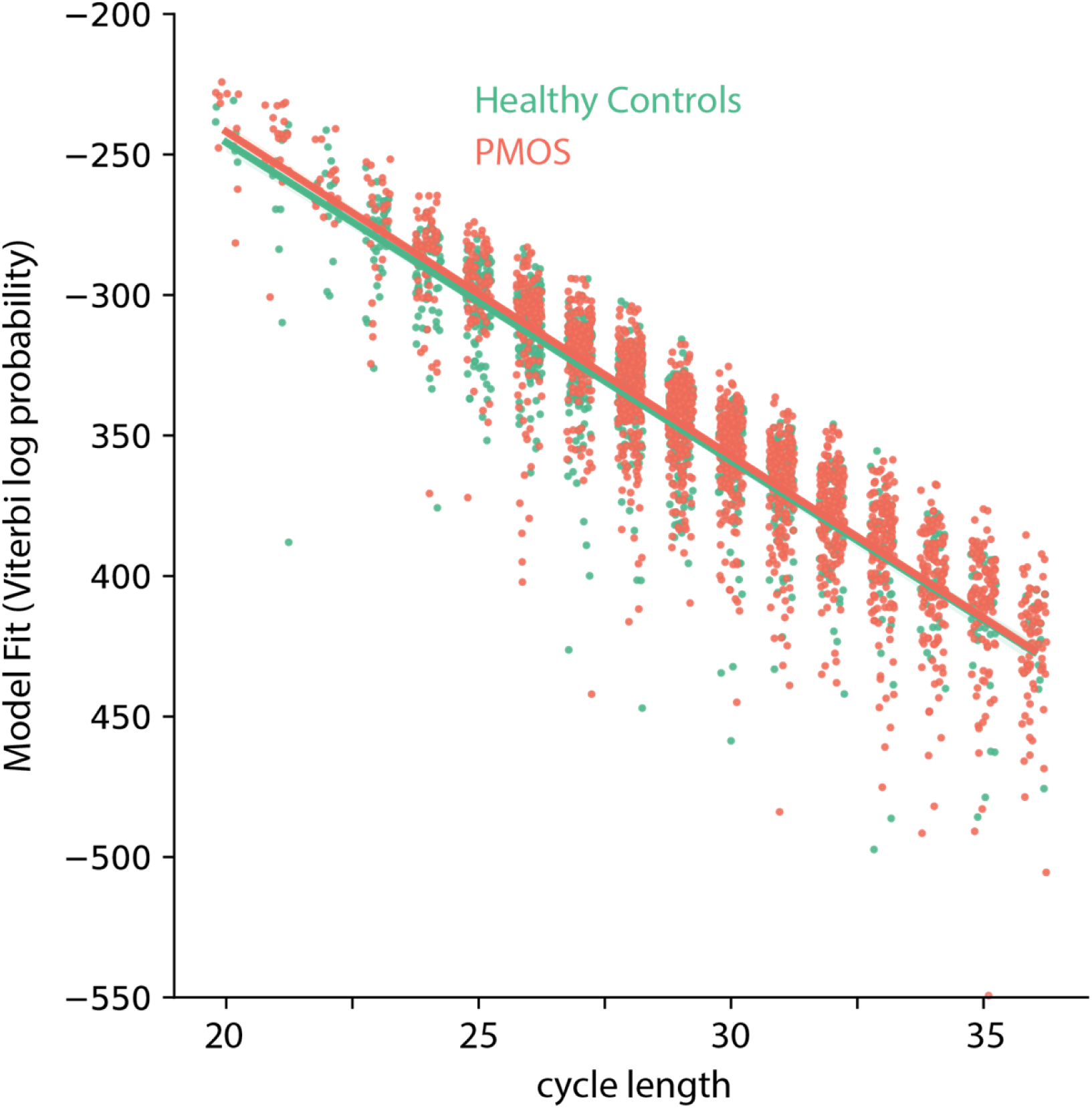
**arHMM fit to cycle data depends on cycle length but not PMOS status**: We use the viterbi algorithm to calculate a cycle-based probability of the observed hormone levels and the most probable path through the phase states. We use this as a measure of model fit. Longer cycles have lower probabilities, as is expected because of how this probability is calculated. The scatterplot above (with x-jitter to better visualize the probabilities for different cycle lengths) along with best-fit lines for healthy controls and individuals with PMOS illustrates that the arHMM does not differ in its ability to fit the cycles of these two populations.

## Notes

### Competing Interest Statement

The authors have declared no competing interest.

### Funding Statement

Yes

### Author Declarations

The data is stored according to the consent given by users in the data use agreement. This study (protocol 2023/10/11) has been verified by the Institutional Review Board at Duquesne University as Exempt according to 45CFR46.101(b)(4): (4) Secondary Research Uses of Data or Specimens on 10/21/2023.

